# Treatment Success and associated factors among patients with Pulmonary Tuberculosis attending Kampala Capital City Authority health facilities: A retrospective cohort study

**DOI:** 10.1101/2022.12.20.22283758

**Authors:** Cathbert Tumusiime, Achilles Katamba, Lydia Nakiyingi, Joan Kalyango

## Abstract

**Background:** TB treatment success remains low in Uganda at 82%, below the recommended WHO target (≥90%). Consequences of poor treatment outcome include; increased MDR-TB prevalence, treatment costs and death. Kampala Capital City Authority (KCCA) public health facilities are congested which compromises the care given to pulmonary tuberculosis patients (PTB) that affects the treatment success of patients. However, there is scarce information regarding factors that are associated with treatment success among PTB patients in KCCA public health facilities.

**General Objective:** To determine the treatment success and associated factors among patients with pulmonary tuberculosis attending KCCA public health facilities in Kampala between July 2019 and June 2020.

**Methods:** A retrospective cohort study that involved review of records for 772 PTB patients who were enrolled on TB treatment in five KCCA health facilities from July 2019 to June 2020. Data on socio-demographic and clinical factors was abstracted from health facility TB registers. Data was entered in epidata and analyzed using STATA_v14 software. A modified poison regression model with robust standard errors was used in analysis and risk ratios were reported.

**Results:** Treatment success was 87.2% (CI: 84.2%-89.1%), PTB patients who cured accounted for 413 (53.5%) and 260 (33.7%) completed treatment. Factors associated with PTB treatment success were: being classified as a clinically diagnosed PTB patients (aRR= 0.8, CI: 0.53 - 0.94, P value =0.021) and having a positive HIV/AIDS status (aRR= 0.7, CI: 0.43 - 0.88, P value =0.006) reduced treatment success and having a community volunteer as a treatment supporter was associated with increased treatment success (aRR= 1.2, CI: 1.06 - 3.28, P value =0.028).

**Conclusion:** Over 80% of PTB patients in KCCA public health facilities achieve treatment success although this is still below the WHO target. Factors associated with TB treatment success include; being classified as a clinically diagnosed PTB patient, having a positive HIV/AIDS status as factors that reduce treatment success and having a community volunteer as a treatment supporter improves treatment success. Efforts such as consistent follow-ups should be encouraged among clinically diagnosed and HIV/AIDS positive PTB patients. Additionally, community volunteers should be empowered to support PTB patients.

## Background

Tuberculosis (TB) is one of the top 10 causes of mortality and the leading cause from a single infectious agent above Human Immunodeficiency Virus/Acquired Immunodeficiency Syndrome (HIV/AIDS). TB control is based on the 3 pillars (integrated patient centered care and prevention; bold policies and supportive systems and intensified research and innovation) of the End TB by 2030 strategy of the United Nations Sustainable Development Goals (SDGs). The management and control of pulmonary tuberculosis (PTB) is important not only to monitor for resistance but also to check for severity, treatment response, limit its spread and improve treatment success (United Nations, 2019).

In 2019, a total of 1.4 million people died from TB including 208,000 (14.9%) people with HIV/AIDS. Worldwide an estimated 10.4 million people fell ill with TB whereby; 5.6 million were men, 3.2 million were women and 1.2 million were children. Of these new TB cases, the largest number occurred in the World Health Organization (WHO) South-East Asian region with 44%, followed by the WHO African region, with 25% (WHO, 2018b). Additionally, 57% of pulmonary tuberculosis cases in 2019 were bacteriologically confirmed, a slight increase of about 2% from 55% recorded in the year 2018 (WHO, 2020).

Sub-Saharan Africa has the second highest burden of TB and the slowest decline in the number of TB incident cases (WHO, 2018a), with a low TB treatment success. According to WHO, the treatment success rate among persons with PTB in sub–Saharan Africa is 82% and is below the WHO recommended treatment success rate of at least 90%. Uganda is one of the 30 WHO-designated countries with a high burden of TB/HIV co-infection (WHO, 2021).

Tuberculosis remains a health emergency and given that significant resources are now being diverted to Corona Virus Disease Nineteen (COVID-19) pandemic management, it is important not to lose sight of the goal of tuberculosis control as this will threaten important milestones, gains and ambitions towards achieving the set treatment success targets (WHO, 2020).

According to the National TB and Leprosy Control Programme (NTLP), during the period July 2019 to June 2020, the Treatment Success Rate (TSR) was 77.9%, this was a slight increase from 76.3% for July 2018 to June 2019 but this is still below the expected national target and global target of 90% (WHO, 2020). KCCA public health facilities play a significant role in provision of PTB management services given the current treatment TB programs being implemented in these facilities. This low treatment success has a potential of increasing drug-resistant TB, TB treatment costs and death if not addressed.

In Kampala, there are eight (8) public health facilities that are directly managed by KCCA with five (5) of these 8 public health facilities running active TB diagnosis and treatment units. Kampala serves a dynamic population that extends beyond the planned numbers in the catchment which makes planning hard especially in handling of drug stockouts (KCCA, 2020). KCCA public health facilities are congested which might compromise the care given to PTB patients that can reduce chances successful treatment (KCCA, 2020). That notwithstanding, there is inadequate information about the factors associated with treatment success among pulmonary PTB patients seeking treatment services from Kampala public health facilities. The study aimed to determine the treatment success and associated factors among patients with pulmonary tuberculosis attending Kampala Capital City Authority health facilities. The findings provide knowledge on factors associated with tuberculosis treatment success in Kampala that can help to inform targeted interventions and policy reviews for refining modalities of TB care in public health facilities in Uganda.

## Methods

### Study setting

This study was done in Kampala, which is the capital city of Uganda. In the capital, there are eight (08) KCCA public health facilities; Kisenyi HC IV, Kawaala HC IV, Kisugu HC III, Kiswa HC III, Komamboga HC III, Kitebi HC II, Bukoto HC II and City Hall clinic HC II. Only the first five facilities have functional TB diagnosis and treatment units.

### Study design

This was a retrospective cohort review of standard TB register (HMIS TB 009) of pulmonary PTB patients who were enrolled on TB treatment during the period from July 2019 to June 2020.

### Population

This comprised all PTB patients who sought treatment at KCCA public health facilities with TB diagnosis and treatment units in Kampala, Uganda. PTB patients who sought treatment at KCCA directly managed health facilities for the cohort the period from July 2019 to June 2020 whose data was accessible. PTB patients aged 15 years and above whose data was available within health unit TB registers for the period of July 2019 to June 2020 and met the eligibility criteria.

### Sample size determination

The sample size for objective 1 was calculated using a single population formula with 95% confidence interval (CI), a 5% margin of error using the (Kish, 1965). For objective 2, we computed a sample size for two proportions based on proportions in previous studies (Chaves et al., 2019). Then we took the largest sample size (768) which was obtained in objective 1.

### Data collection

The data were collected through reviewing the necessary documents (HMIS TB 009) of the TB patients using a pre-tested structured data extraction format which was developed by considering the variables to be studied. The format contained all the important sociodemographic and clinical characteristics. Three (3) research assistants working as data clerks in the TB units were trained for one day and gathered the data. Data was collected from 772 patient records with four records above the calculated sample size. The filled forms were checked for completeness daily during the data collection. The whole process was supervised by the principal investigator.

### Data management and statistical analysis

Data abstracted from health unit register (HMIS TB 009) was entered through epidata (v4.6.0.5) and analyzed using STATA (v14). Frequencies, proportions were used to describe the study participants in relation to socio-demographic and clinical characteristics. For objective one, the treatment success for pulmonary TB was obtained by computing the proportion of pulmonary tuberculosis patients cured and those who completed treatment in the study period and was reported with the corresponding adjusted confidence interval. To answer objective two, factors associated with PTB treatment success were established through a regression analysis of the dependent variable against each of the independent variables. Data was adjusted for clustering at health facility level by way of declaring it as a survey dataset to assess for similarity of TB patients from each of the selected facilities and a negligible design effect of 1.1 was found. Known risk factors to PTB treatment success like HIV/AIDS status were maintained throughout analysis irrespective of statistical significance. Also, data was assessed for interaction (>20%) and confounding (>10%). A variable that had a p-value ≤ 0.2 in the bivariate analysis was entered into the multivariate regression model.

A modified Poisson regression analysis model with robust standard errors was used to perform multivariate analysis for all statistically significant variables identified at the bivariate analysis and reported the results as risk ratios (RR). Each RR was reported with the corresponding 95% confidence interval (CI), for both the unadjusted and adjusted results. The data analysis was performed at the 5% level of significance and a deviance goodness of fit test was conducted.

## Results

### Study profile of persons with PTB in KCCA public health facilities

Of 3,004 persons with TB enrolled on TB treatment during the cohort of July 2019 to June 2020 (figure 2), the study excluded the following: 702 patients with extra pulmonary and MDR TB, 847 patients who were aged below 15 years and 683 who missed information on the critical variables of interest. All the records (772) that remained relevant were then abstracted and entered for subsequent analyses over the computed sample size of 768 since this was an additional four (4) patient records.

**Figure 1:**
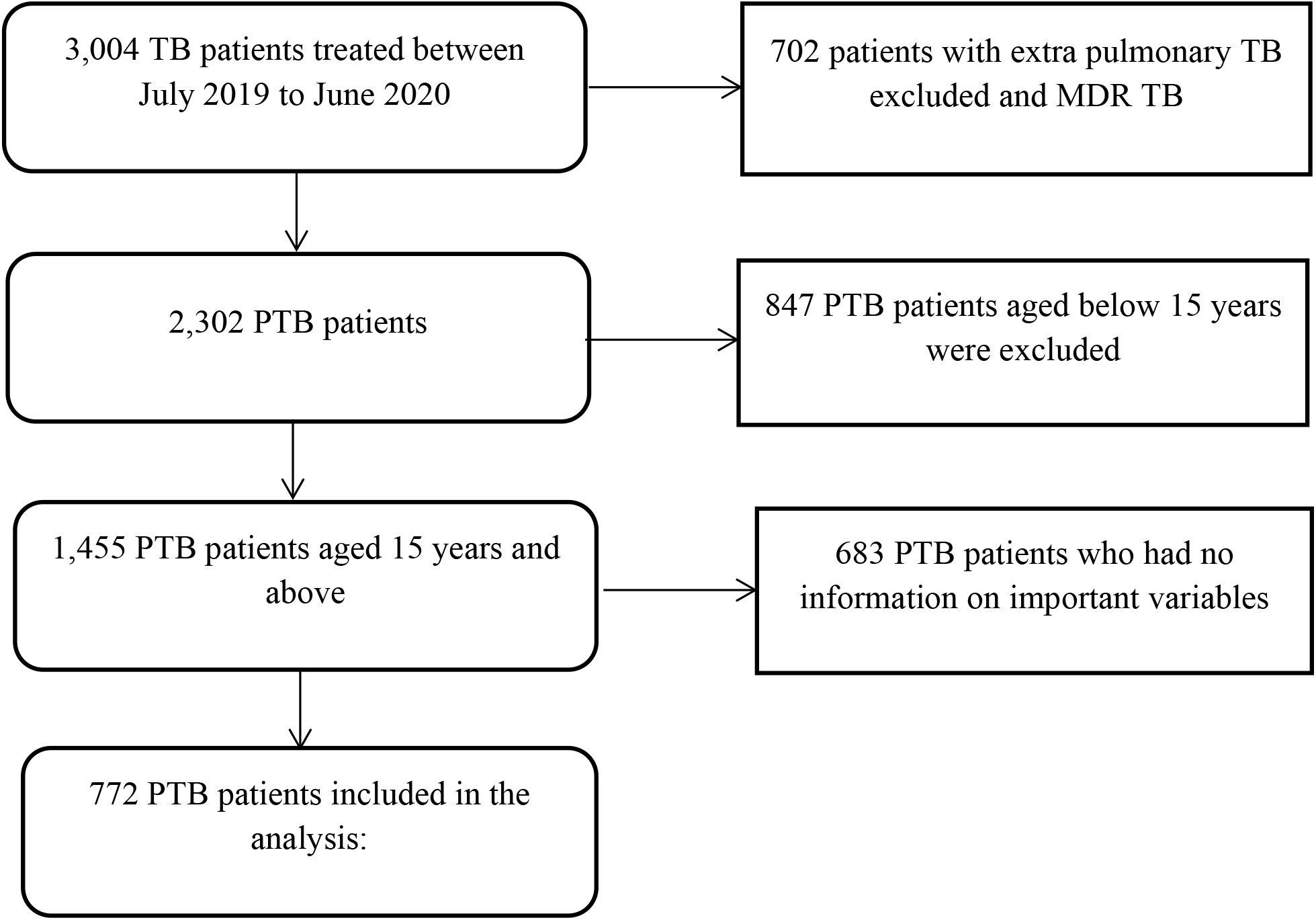
Study profile of persons with PTB in KCCA public health facilities

**Figure 2:**
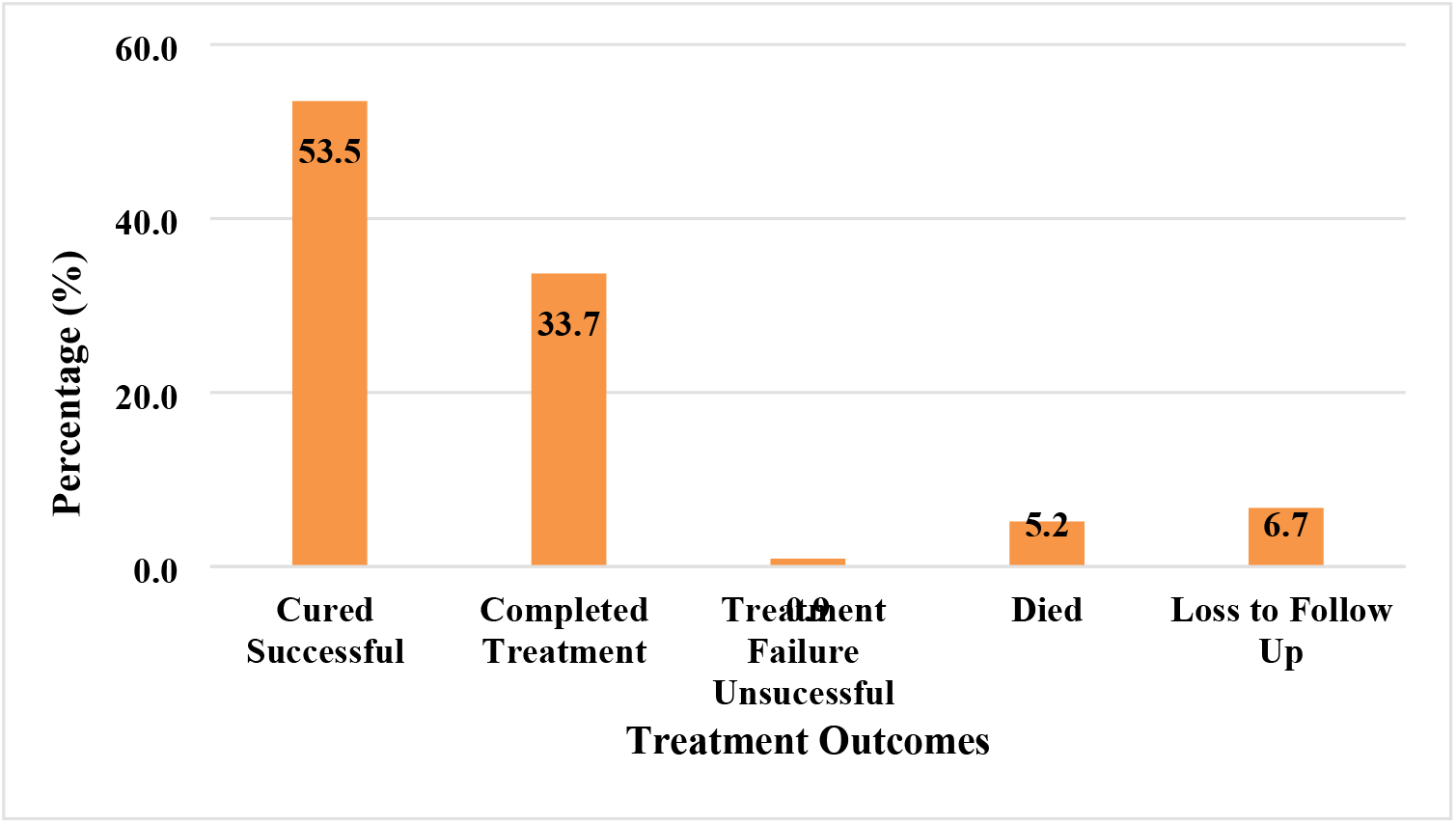
Distribution of treatment outcomes of 772 patients at the KCCA public health facilities. **Source: *Field data; November/December 2021***

### Socio-demographic and clinical characteristics of the 772 PTB patients

#### Socio-demographic characteristics

A total of 772 participant records were abstracted from health facility HMIS TB 009 registers from five (05) KCCA health facilities. The mean age (standard deviation) was 33.6 years (11.4). Age of the patients was then categorized based on a PTB study by (Thomsen et al., 2017). Majority (67.6%, n=521) were males. More than half of the patients (56.8%, n=231) had a normal weight and over 75.7% (n=584) were residents of Kampala. Of those who had a risk group classification, most (92.7%, n=290) were contacts to previous TB patients. Table 1 summarizes.

**Table 1:** Socio-demographic characteristics of PTB patients in the KCCA public health facilities (n=772)

| Characteristic | Frequency (n) | Percent (%) |
| --- | --- | --- |
| **Age** |  |  |
| 15-24 | 172 | 22.2 |
| 25-44 | 468 | 60.8 |
| 45-64 | 117 | 15.1 |
| ≥ 65 | 15 | 1.9 |
| **Sex** |  |  |
| Male | 522 | 67.6 |
| Female | 250 | 32.4 |
| **BMI (n=407)** |  |  |
| Under weight (<18.5) | 121 | 29.7 |
| Normal weight (18.5-24.9) | 231 | 56.8 |
| Overweight (25-29.9) | 22 | 8.1 |
| Obese (≥35) | 33 | 5.4 |
| **Residence (parish)** |  |  |
| Outside Kampala | 188 | 24.3 |
| Within Kampala | 584 | 75.7 |
| **Risk Group (n=313)** |  |  |
| Health Worker | 02 | 0.6 |
| TB Contact | 290 | 92.7 |
| Uniformed personnel | 05 | 1.6 |
| Prisoner | 04 | 1.2 |
| Others** | 12 | 3.9 |

\*\* These included hawkers, teacher, carpenter, driver, cleaner and hotelier.
**Source: Field data; November/December 2021**

### Clinical Characteristics

Results indicated that majority (60.5%, n=467) of the patients were bacteriologically confirmed pulmonary (BC PTB) patients. Over 90% (n = 729) were newly registered cases, a considerable proportion (34.3%) of the patients were also co-infected with HIV/AIDS whereas 16.4% (n=108) were found to be diabetic at the time of TB diagnosis. Nutrition was assessed using Mid-Upper Arm Circumference (MUAC) and it was established that majority (61.0%, n=175) were normal. Nearly 50% (n=381) of the patients were on Digital Community Directly Observed Therapy (DC-DOT) and majority (89.9%, n=694) of the patients reported that they were being supported by a family member. Table 2 shows the clinical characteristics of the patients.

**Table 2:** Clinical characteristics of PTB patients in the KCCA public health facilities (n=772)

| Characteristic | Frequency (n) | Percent (%) |
| --- | --- | --- |
| **Disease classification** |  |  |
| BC PTB<sup>1</sup> | 467 | 60.5 |
| CD PTB<sup>2</sup> | 305 | 39.5 |
| **Type of TB patient** |  |  |
| New | 729 | 94.4 |
| Relapse | 36 | 4.7 |
| Treatment Failure | 07 | 0.9 |
| **HIV/AIDS Status** |  |  |
| Positive | 265 | 34.3 |
| Negative | 507 | 65.7 |
| **DM status (n=659)** |  |  |
| DM+ | 108 | 16.4 |
| DM- | 551 | 83.6 |
| **MUAC (n=287)** |  |  |
| Normal ($\geq 22.5$ cm) | 175 | 61.0 |
| Malnourished ($< 22.5$ cm) | 112 | 39.0 |
| **Treatment model** |  |  |
| Facility DOT | 32 | 4.1 |
| Digital Community DOT | 381 | 49.4 |
| Non-Digital Community DOT | 359 | 46.5 |
| **Treatment supporter** |  |  |
| Community volunteer | 78 | 10.1 |
| Family Member | 694 | 89.9 |

**Source: Field data; November/December 2021**

### Treatment success Rate of 772 patients in KCCA public health facilities

Overall, results showed that treatment success was 87.2% (CI: 81.8% - 89.1%) [where cured – 413(53.5%) and completed treatment – 260(33.7%)]. Figure 3 shows the distribution of treatment outcomes of study patients in KCCA public health facilities.

### Bivariate analysis of factors associated with PTB treatment success among 772 patients in KCCA public health facilities

At bivariate analysis level, the independent variables that were significantly associated with PTB treatment success were sex (cRR=0.7, P value=0.147), being classified as a clinically diagnosed PTB patient (cRR=0.6, P value=0.010), being HIV/AIDS positive (cRR=0.7, P value= 0.002), being on a facility DOT treatment model (cRR=0.4, P value = 0.135), being supported by a community volunteer to take tuberculosis drugs (cRR=1.3, P value=0.015). Results of bivariate analysis of factors associated with PTB treatment success are summarised in Table(s) 3a and b.

**Table 3a:** Bi variate analysis of socio-demographic factors associated with treatment success among PTB patients in KCCA public health facilities (n=772)

| Characteristic | Treatment Success |  | cRR(95% CI) | P-value |
| --- | --- | --- | --- | --- |
|  | successful<br>n(%) | unsuccessful<br>n(%) |  |  |
| **Age** |  |  |  |  |
| 15-24 | 153 (89.9) | 19 (11.1) | 1 |  |
| 25-44 | 407(87.0) | 61(13.0) | 2.2(0.75 4.06) | 0.915 |
| 45-64 | 100(85.4) | 17(14.6) | 0.6(0.41 3.52) | 0.776 |
| ≥ 65 | 13(86.7) | 2(13.3) | 0.4(0.12 8.13) | 0.894 |
| **Sex** |  |  |  |  |
| Male | 460 (88.1) | 62(11.9) | 1 |  |
| Female | 213 (85.2) | 37 (14.8) | 0.7(0.36 0.93) | **0.147** |
| **BMI** |  |  |  |  |
| Under weight (<18.5) | 107(88.4) | 14(11.6) | 1 |  |
| Normal weight (18.5-24.9) | 204(88.3) | 27(11.7) | 1.1(0.64 1.58) | 0.974 |
| Overweight (25-29.9) | 18(81.8) | 4(18.2) | 0.3(0.19 2.34) | 0.515 |
| Obese (≥35) | 29 (87.9) | 4 (12.1) | 0.5(0.21 9.28) | 0.274 |
| **Residence (parish)** |  |  |  |  |
| Within Kampala | 511(87.6) | 72(12.4) | 1 |  |
| Outside Kampala | 160(85.1) | 28(14.9) | 0.9(0.68 2.63) | 0.402 |

**Table 3b:**
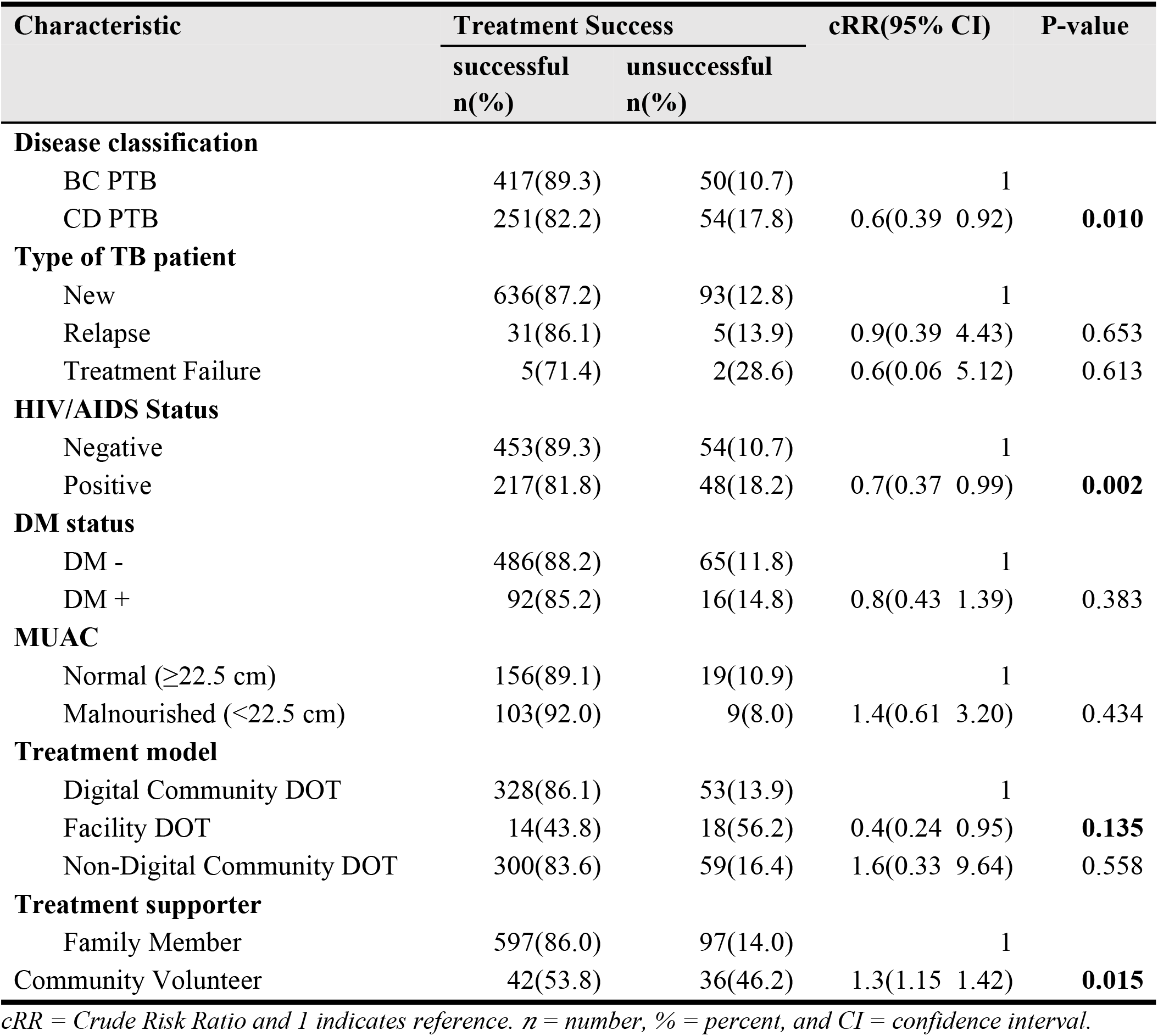
Bi variate analysis of clinical factors associated with treatment success among PTB patients in KCCA public health facilities (n=772)

### Multivariate analysis of factors associated with PTB treatment success among 772 patients in KCCA public health facilities

After adjusting for other covariates, factors significantly associated with PTB treatment success among 772 patients studied were: being classified as a clinically diagnosed PTB patient (aRR= 0.8, CI: 0.53 - 0.94, P value =0.021), having a positive HIV/AIDS status (aRR= 0.7, CI: 0.43 - 0.88, P value =0.006) as factors responsible for reduced treatment success. Results also indicted that having a community volunteer as a treatment supporter was significantly associated with increased treatment success of the PTB patient (aRR= 1.2, CI: 1.06 - 3.28, P value =0.028). There was neither interaction nor confounding with any of significant variables. Results of a multivariate analysis of factors associated with treatment success of PTB patients in KCCA public health facilities are presented in table 4.

**Table 4:**
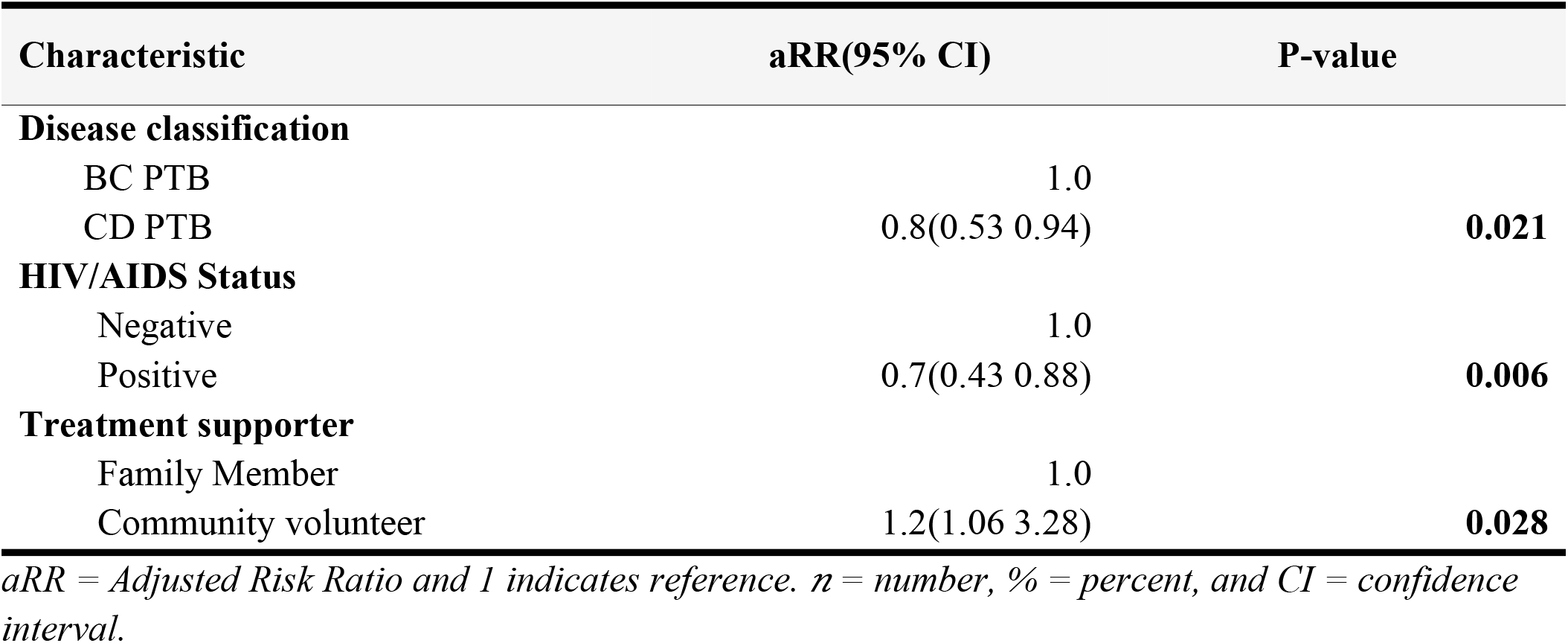
Multivariate analysis of factors associated with PTB treatment success among 772 patients in KCCA public health facilities.

| Characteristic | aRR(95% CI) | P-value |
| --- | --- | --- |
| **Disease classification** |  |  |
| BC PTB | 1.0 |  |
| CD PTB | 0.8(0.53 0.94) | **0.021** |
| **HIV/AIDS Status** |  |  |
| Negative | 1.0 |  |
| Positive | 0.7(0.43 0.88) | **0.006** |
| **Treatment supporter** |  |  |
| Family Member | 1.0 |  |
| Community volunteer | 1.2(1.06 3.28) | **0.028** |

*aRR = Adjusted Risk Ratio and 1 indicates reference. n = number, % = percent, and CI = confidence interval.*

## Discussion

### Treatment success among 772 pulmonary tuberculosis patients in 05 KCCA public health facilities

During the study period, the treatment success rate of 772 pulmonary tuberculosis patients who sought TB treatment services from the five (5) KCCA public health facilities between July 2019 to June 2020 was 87.2% (CI:81.8%,89.1%). Thus, approximately 87 PTB patients in every 100 patients diagnosied with PTB have a successful treatment outcome at the end of the treatment period. According to the Ministry of Health, this treatment success rate is above the treatment success rate for Uganda which stands at 82%. Though majority of the patients responded to the treatment, the rate of treatment success was below the national target by the Ministry of Health target of ≥90% (MOH, 2017).

PTB treatment success in KCCA public health facilities is higher than that in Kampala. Our findings revealed a higher treatment succes than that in a study done in Ghana which found treatment success at 68.5% (Agyare et al., 2021). Another study done in sub-saharan Africa revealed a treatment success rate of 76.2% among pulonary tuberculosis patients and this was lower than that found out in this study (Izudi, Semakula, Sennono, Tamwesigire, & Bajunirwe, 2019).

Furthermore, lower treatment success rates were seen in a study done in Morocco (53.6%) (El Hamdouni et al., 2019), Eastern Ethiopia (81%) (Zenebe & Tefera, 2016) and India (81%) (Madan et al., 2018). Our findings were however, lower than those of a study conducted in China which established the treatment success of pulmonary tuberculosis at 95.0% (Wen et al., 2018).

A possible explanation for the difference in the results could be there are TB programmes being implemented towards improving TB treatment success. Another reason could be due to the fact that this study was conducted in only public health facilities. The implication here is that there is a likelihood of patients developing multi-drug resistant TB, high costs associated with TB management

### Factors associated with TB treatment success of pulmonary tuberculosis among 772 patients in 05 KCCA health facilities

Our findings showed that disease classification was significantly associated with PTB treatment success. Thus, clinically diagnozed PTB patients were 20% less likely to have successful treatment compared to bacteriologically confirmed PTB patients (aRR =0.8, P-value = 0.021; CI: 0.53 - 0.94). Our findings are inconsistent with a study done in Kenya where treatment success was nearly similar between clinically diagnozed and bacteriologically confirmed PTB patients but the mortality rate was 5 times higher among clinically diagnosed patients (Abdullahi, Moses, Sanga, & Annie, 2021).

A possible explanation could be misdiagnosis of other conditions that mimic TB as TB, thus the unsuccessful outcome like mortality.

Patients’ HIV/AIDS status was significantly associated with PTB treatment success. Patients who were HIV/AIDS negative had a 30% more chance of experiencing treatment success as compared to those patients who were HIV/AIDS positive (aRR =0.7, P-value = 0.006; CI: 0.43 - 0.88). These findings are in line with a study done in Peru (Velásquez et al., 2019), which demonstrated that positive HIV status was a strong predictor of unsuccessful outcome among patients treated for tuberculosis. Many other studies have found HIV/AIDS to be a known risk factor to TB treatment outcome among PTB patients. A possible explanation could be that there are other comorbidities in addition to the TB in HIV such as severe disease, delayed diagnosis, other drugs causing drug-drug interactions and side effects.

Type of treatment supporter was significantly associated with treatment success of PTB patients. Patients whose type of treatment supporter was a community volunteer had about 20% chances of achieving treatment success compared to PTB patients whose treatment supporter was a family member (aRR =1.2, P-value = 0.028; CI: 1.06 - 3.28).

These findings are similar to those from a study on type of treatment supporters in successful completion of PTB treatment by (Hussain et al., 2018) where treatment success rate of the TB patients was high among those who were supported by a family member or community volunteer compared to patients who had no treatment supporter. A possible explanation could be that supervision of persons with PTB by a community volunteer or family member promotes adherence to medical advice and thus good treatment outcomes which ultimately improve the treatment success.

Patients’ treatment model was not associated with PTB treatment success. However, at bivariate level, patients who were on Facility DOT were 60% less likely to get a successful outcome compared to patients who were on Digital Community DOT. These findings are in agreement with a meta-analysis study by (Zhang, Ehiri, Yang, Tang, & Li, 2016), where PTB cases demonstrated that digital community-DOT promoted successful treatment compared with facility DOT treatment. A possible explanation could be that the patients who are on Facility DOT maybe constrained by frequent travels to and from the facility for drugs which may have financial implication with regard to transportation, resulting into missing out on drug days thus affecting treatment success.

Mid-Upper Arm Circumference was not associated with treatment success among PTB patients. Our findings are in agreement with a study done in the Philippines (Lee et al., 2019) in which MUAC was not associated with treatment success in the group where patients either had a normal MUAC or were malnourished. This could be because patients in KCCA public health facilities are always given health education prior to treatment and encouraged to feed well once diagnosed with TB and thus there are minimal chances of being malnourished.

Patients body mass index (BMI) was not associated with treatment success. These findings are contrary to those of a study done in Uganda (Mupere et al., 2014), which found that BMI measurements were associated with the outcome in the initial three months of tuberculosis treatment of the patient. A possible explanation could be that this study did not have sufficient power to detect the influence of BMI on treatment success due to missing information that is not adequately captured in the HMIS tools.

Patients’ diabetes mellitus (DM) status was also not associated with treatment success. Our findings were consistent with a study done Netherlands (Bates, Marais, & Zumla, 2015) which found a negative impact of diabetes on treatment success. However, our findings were inconsistent with a study done in Kelantan, Malaysia by (Ahmad et al., 2020) which found that success rate in TB patients with diabetes mellitus was higher compared to TB patients without diabetes mellitus. This might be explained by delayed clearance resulting in routine switching to a two-drug regimen in patients who are still culture-positive or lowered rifampicin levels in diabetes patients.

Age was not associated with PTB treatment success. These findings are in line with a study done in China (Ai et al., 2010) which found that age of the patient was not significantly associated with treatment success. According to (Ncube et al., 2017), the elderly have the worst outcomes among all the age groups which may be related to immunosuppressant comorbidities or other age-related diseases mis-classified as TB. However, our study did not have sufficient power to detect the influence of age on treatment success among different age groups especially those aged ≥65.

The study findings did not show association between sex and pulmonary TB treatment success. However, at bivariate level, male PTB patients had 30% more chances to achieve treatment success as compared to the female counterparts. These findings are similar with other studies in Uganda (Nakanwagi-Mukwaya et al., 2013) where sex of the patient had no significant influence on the treatment outcome whether as successful or unsuccessful at multivariate level. A possible explanation could be that the TB treatment service seeking behavior for females slightly varies from that of males, but the variation is not strong enough to influence treatment success.

Residential status was also not associated with PTB treatment success. Our findings are contrary to those from a study done in Uganda (Robsky et al., 2020) which found a protective association between longer distance from home to chosen treatment facility and treatment success. This could be because the KCCA public health facilities are few in Kampala with free services and thus whether a patient comes from near or far, they find it convenient to access free treatment services.

## Limitations of the study

There are some limitations that were faced in the course of conducting this study and these include;

i. Use of retrospective secondary data where by some important variables which might have impact on treatment outcome of PTB patients, like socioeconomic characteristics (income, family size, educational status, social support, distance to the health facility) as well as behavioral factors (knowledge and attitude about the diseases, alcohol abuse, cigarette smoking, illicit drug use) were not recorded.
ii. There was missing information on some variables which could have had a bearing on the overall results since 683 patient records with missing important information were omitted. However, these were similar to the ones retained during the study on important demographic characteristics.
iii. This study also involved only patients in public hospitals. Thus, the findings may not be very applicable to the patients in the private setting.

## Conclusions and Recommendations

### Conclusions

- About 87 in every 100 (CI: 0.84 0.89) PTB patients in KCCA public health facilities are likely to achieve treatment success. This is, however, lower than the national target (≥90%).
- Being classified as a clinically diagnosed pulmonary tuberculosis patient, being HIV/AIDS positive and being supported by a community volunteer during treatment were found to be statistically significant associated with PTB treatment success.

### Recommendations

#### KCCA

- KCCA public health facilities TB DTUs should give adequate attention to clinically diagnosed PTB patients as these have an influence in achieving treatment success.
- Emphasis should be put on completion of data capture of all indicators in the TB register to mitigate the information loss.

### Ministry of Health

Community volunteers should be empowered to support PTB patients in order to improve treatment success.

### Academia

Other studies could be carried out to assess the influence of other factors like socio-economic and behavioral characteristics on PTB patients’ treatment success in public health facilities.

## Data Availability

The datasets of the study are available on reasonable request from the corresponding author.

## List of abbreviations

AIDS: Acquired Immunodeficiency Syndrome
aRR: Adjusted Risk Ratio
ART: Anti-Retroviral Treatment
BC: PTB Bacteriologically Confirmed Pulmonary Tuberculosis
BMI: Body Mass Index
CBD: Central Business District
CD: PTB Clinically Diagnosed Pulmonary Tuberculosis
CDC: Centers for Disease Control and Prevention
CHS: College of Health Sciences
CI: Confidence Interval
COVID-19: Corona Virus Disease Nineteen
cRR: Crude Risk Ratio
DM: Diabetes Mellitus
DOT: Directly Observed Treatment
DTU: Diagnosis and Treatment Unit
HIV: Human Immunodeficiency Virus
HMIS: Health Management Information System
HSDP: Health Sector Development Plan
INH: Isoniazid
IRB: Institutional Review Board
KCCA: Kampala Capital City Authority
LTF: Loss to Follow-up
MDR-TB: Multi-Drug Resistant Tuberculosis
MOH: Ministry of Health
MUAC: Mid-Upper Arm Circumference
NTLP: National Tuberculosis & Leprosy Control Programme
PTB: Pulmonary Tuberculosis
SDGs: Sustainable Development Goals
SOMREC: School of Medicine Research Ethics Committee
TSR: Treatment Success Rate
UN: United nations
WHO: World Health Organization

## Declarations

### Ethical consideration

Permission to conduct the study was obtained from Clinical Epidemiology Unit (CEU). Institutional Review Board (IRB) approval to conduct the study was sought from School of Medicine Research and Ethics Committee (SOMREC) under reference number: MAK-SOMREC-2021-182. The researcher sought for waiver of participant consent from SOM-REC since data was abstracted from patient records and thus no direct contact with study participants. Permission was obtained from the Directorate of Public Health and Environment – Kampala Capital City Authority (KCCA). In addition, permission was sought from the facility in-charge(s) for each of the selected health facilities prior to accessing the patient records. Patient identification numbers were used instead of names and the data collected was safely kept under lock and key and on password protected computers with restricted access for only purposes of this research while ensuring confidentiality of all study related data.

### Consent for publication

Not applicable.

### Availability of data

The datasets of the study are available on reasonable request from the corresponding author.

### Authors contributions

All authors were involved in the conception of the study, design, data acquisition, data analysis and interpretation. The manuscript was also developed through active participation of all authors.

## Acknowledgements

Administration of Kampala Capital City Authority (KCCA) specifically the Directorate of Public Health and Environment (DPHE), the health facility in-charges for Kisenyi HC IV, Kawaala HC IV, Kiswa HC III, Kisugu HC III, and Komamboga HC III, research assistants, my parents, and my classmates. Finally, I am very thankful to the Lord God Almighty for giving me wisdom, knowledge, favor and making this work possible.

## Authors’ information

CT is a candidate for Master of Science in Clinical Epidemiology and Biostatistics at the Clinical Epidemiology Unity, School of Medicine, College of Health Sciences, Makerere University.

AK is a senior lecturer in the in Clinical Epidemiology and Biostatistics at the Clinical Epidemiology Unity, School of Medicine, College of Health Sciences, Makerere University

LN is a Senior Lecturer and Physician (Infectious Diseases), Department of Medicine, Makerere College of Health Sciences, Makerere University.

JK is an associate professor at the Clinical Epidemiology Unity, School of Medicine, College of Health Sciences, Makerere University.

## References

Abdullahi, O., Moses, N., Sanga, D., & Annie, W. (2021). The effect of empirical and laboratory-confirmed tuberculosis on treatment outcomes. Scientific Reports, 11(1), 14854. doi:10.1038/s41598-021-94153-0

Agyare, S. A., Osei, F. A., Odoom, S. F., Mensah, N. K., Amanor, E., Martyn-Dickens, C., … Yeboah, E. O. (2021). Treatment Outcomes and Associated Factors in Tuberculosis Patients at Atwima Nwabiagya District, Ashanti Region, Ghana: A Ten-Year Retrospective Study. Tuberculosis Research and Treatment, 2021, 9952806. doi:10.1155/2021/9952806

Ahmad, S. R., Yaacob, N. A., Jaeb, M. Z., Hussin, Z., Wan, M. W., & Mohd, Z. (2020). Effect of Diabetes Mellitus on Tuberculosis Treatment Outcomes among Tuberculosis Patients in Kelantan, Malaysia. Iranian journal of public health, 49(8), 1485–1493. doi:10.18502/ijph.v49i8.3892

Ai, X., Men, K., Guo, L., Zhang, T., Zhao, Y., Sun, X., … van den Hof, S. (2010). Factors associated with low cure rate of tuberculosis in remote poor areas of Shaanxi Province, China: a case control study. BMC public health, 10(1), 112. doi:10.1186/1471-2458-10-112

Bates, M., Marais, B. J., & Zumla, A. (2015). Tuberculosis Comorbidity with Communicable and Noncommunicable Diseases. Cold Spring Harbor perspectives in medicine, 5(11), a017889. doi:10.1101/cshperspect.a017889

Chaves, T., Ninfa, M., Quijano, R. J. J., Porras, A. P. S., Arriaga, M. B., Netto, E., & Martins. (2019). Factors predictive of the success of tuberculosis treatment: A systematic review with meta-analysis. PLOS ONE, 14(12), e0226507–e0226507. doi:10.1371/journal.pone.0226507

El Hamdouni, M., Bourkadi, J. E., Benamor, J., Hassar, M., Cherrah, Y., & Ahid, S. (2019). Treatment outcomes of drug resistant tuberculosis patients in Morocco: multi-centric prospective study. BMC Infectious Diseases, 19(1), 316. doi:10.1186/s12879-019-3931-5

Hussain, S., Hasnain, J., Hussain, Z., Badshah, M., Siddique, H., Fiske, C., & Pettit, A. (2018). Type of Treatment Supporters in Successful Completion of Tuberculosis Treatment: A Retrospective Cohort Study in Pakistan. The open infectious diseases journal, 10, 37–42. doi:10.2174/1874279301810010037

Izudi, J., Semakula, D., Sennono, R., Tamwesigire, I. K., & Bajunirwe, F. (2019). Treatment success rate among adult pulmonary tuberculosis patients in sub-Saharan Africa: a systematic review and meta-analysis. BMJ Open, 9(9), e029400–e029400. doi:10.1136/bmjopen-2019-029400

KCCA. (2020). Directorate of Public Health and Environment Annual Report: Kampala Capital City Authority (KCCA). 08, 2–3.

Kirigia, J. M., & Muthuri, R. D. K. (2016). Productivity losses associated with tuberculosis deaths in the World Health Organization African region. Infectious Diseases of Poverty, 5(1), 43. doi:10.1186/s40249-016-0138-5

Kish, L. (1965). Survey sampling. New York: John Wiley & Sons.

Lee, N., White, L. V., Marin, F. P., Saludar, N. R., Solante, M. B., Tactacan-Abrenica, R. J. C., … Cox, S. E. (2019). Mid-upper arm circumference predicts death in adult patients admitted to a TB ward in the Philippines: A prospective cohort study. PLOS ONE, 14(6), e0218193–e0218193. doi:10.1371/journal.pone.0218193

Madan, C., Chopra, K., Satyanarayana, S., Surie, D., Chadha, V., Sachdeva, K. S., … Chauhan, L. (2018). Developing a model to predict unfavourable treatment outcomes in patients with tuberculosis and human immunodeficiency virus co-infection in Delhi, India. PLOS ONE, 13, e0204982. doi:10.1371/journal.pone.0204982

MOH. (2017). Uganda National TB and Leprosy Program July 2016 - June 2017 Report. 14–18.

MOH. (2019). National Tuberculosis and Leprosy Program: National Agenda for Operations and Implementation Research. 15–16.

MOH. (2020). Annual Health Sector Performance Report (AHSPR). Retrieved from Kampala, Uganda:

Mupere, E., Malone, L., Zalwango, S., Okwera, A., Nsereko, M., Tisch, D. J., … Tuberculosis Research Unit at Case Western Reserve, U. (2014). Wasting among Uganda men with pulmonary tuberculosis is associated with linear regain in lean tissue mass during and after treatment in contrast to women with wasting who regain fat tissue mass: prospective cohort study. BMC Infectious Diseases, 14(1), 24. doi:10.1186/1471-2334-14-24

Nakanwagi-Mukwaya, A., Reid, A. J., Fujiwara, P. I., Mugabe, F., Kosgei, R. J., Tayler-Smith, K., … Joloba, M. (2013). Characteristics and treatment outcomes of tuberculosis retreatment cases in three regional hospitals, Uganda. Public health action, 3(2), 149–155. doi:10.5588/pha.12.0105

Ncube, R. T., Takarinda, K. C., Zishiri, C., van den Boogaard, W., Mlilo, N., Chiteve, C., … Sandy, C. (2017). Age-stratified tuberculosis treatment outcomes in Zimbabwe: are we paying attention to the most vulnerable? Public health action, 7(3), 212–217. doi:10.5588/pha.17.0024

Robsky, K. O., Hughes, S., Kityamuwesi, A., Kendall, E. A., Kitonsa, P. J., Dowdy, D. W., & Katamba, A. (2020). Is distance associated with tuberculosis treatment outcomes? A retrospective cohort study in Kampala, Uganda. BMC Infectious Diseases, 20(1), 406. doi:10.1186/s12879-020-05099-z

Thomsen, V. Ø., Zhang, X., Andersen, A. B., Lillebaek, T., Kamper-Jørgensen, Z., Ladefoged, K., … Yang, Z. (2017). Effect of sex, age, and race on the clinical presentation of tuberculosis: a 15-year population-based study. The American journal of tropical medicine and hygiene, 85(2), 285–290. doi:10.4269/ajtmh.2011.10-0630

Tola, A., Minshore, K. M., Ayele, Y., & Mekuria, A. N. (2019). Tuberculosis Treatment Outcomes and Associated Factors among TB Patients Attending Public Hospitals in Harar Town, Eastern Ethiopia: A Five-Year Retrospective Study. Tuberculosis Research and Treatment, 2019, 1503219. doi:10.1155/2019/1503219

United Nations. (2019). The Sustainable Development Goals Report 2019: United Nations.

Velásquez, G. E., Cegielski, J. P., Murray, M. B., Yagui, M. J., Asencios, L. L., Bayona, J. N., … Shin, S. S. (2019). Impact of HIV on mortality among patients treated for tuberculosis in Lima, Peru: a prospective cohort study. (1471-2334 (Electronic)).

Wen, Y., Zhang, Z., Li, X., Xia, D., Ma, J., Dong, Y., & Zhang, X. (2018). Treatment outcomes and factors affecting unsuccessful outcome among new pulmonary smear positive and negative tuberculosis patients in Anqing, China: a retrospective study. BMC Infectious Diseases, 18(1), 104. doi:10.1186/s12879-018-3019-7

WHO. (2018a). Etimates of TB and MDR-TB burden are produced by WHO in consultation with countries. Tuberculosis Profile(2020-02-03).

WHO. (2018b). Global Tuberculosis Report Retrieved from Geneva, Switzerland:

WHO. (2018c). TB Country Profile. Retrieved from https://treattb.org/wp-content/uploads/2018/10/WHO_HQ_Reports-G2-PROD-EXT-TBCountryProfile-uganda.pdf

WHO. (2020). Global Tuberculosis Report Retrieved from Geneva, Switzerland:

WHO. (2021). Global Tuberculosis Report (CC BY-NC-SA 3.0 IGO.). Retrieved from Geneva, Switzerland: https://www.who.int/publications/i/item/9789240037021

Zenebe, T., & Tefera, E. (2016). Tuberculosis treatment outcome and associated factors among smear-positive pulmonary tuberculosis patients in Afar, Eastern Ethiopia: a retrospective study. (1678-4391 (Electronic)).

Zhang, H., Ehiri, J., Yang, H., Tang, S., & Li, Y. (2016). Impact of Community-Based DOT on Tuberculosis Treatment Outcomes: A Systematic Review and Meta-Analysis. PLOS ONE, 11(2), e0147744–e0147744. doi:10.1371/journal.pone.0147744

